# Human brain MRI data of CSF tracer evolution over 72h for data-integrated simulations

**DOI:** 10.1101/2025.07.23.25331971

**Authors:** Jørgen N. Riseth, Timo Koch, Sofie Lysholm Lian, Tryggve Holck Storås, Ludmil T. Zikatanov, Lars Magnus Valnes, Kaja Nordengen, Kent-André Mardal

## Abstract

We present the Gonzo dataset: Brain MRI and derivative data from one healthy male human volunteer (“Gonzo”) before and during the 72 hours after intrathecal injection of the contrast agent gadobutrol into the cerebrospinal fluid (CSF) of the spinal canal. The MRI data records include images highlighting the temporal and spatial evolution of the contrast agent in CSF, brain, and adjacent structures. In addition to raw MRI, we provide derivatives that enable numerical simulations of the transport process under study. Derivatives include T1 maps, tracer concentration maps, diffusion tensor maps, and unstructured triangulated volume meshes of the brain geometry. We also provide brain region markers obtained by image segmentation. A regional statistical analysis of the concentration data complements the image data. The presented data can be used to study the transport behavior and the underlying processes of a tracer in the brain. It is intended to contribute to and inspire new studies on the understanding of tracer transport, method development for image analysis, and simulation of brain fluid transport processes.

## Background & Summary

Cerebrospinal fluid (CSF) surrounds the brain and spinal cord, in the subarachnoid space, and in the ventricular system. The motion of the CSF is dynamic and complex; fluid is secreted in the choroid plexus in the ventricles; the flow of the CSF is also pulsatile with several relevant frequencies driven by heart^1^, lung^2^, vasomotion^3^, and sleep cycles^4^. Apart from acting as a shock absorber for the brain, CSF offers a potential route for drug delivery to the brain, circumventing the blood-brain barrier that hinders most blood-borne substances from reaching the functional brain tissue. CSF is also proposed to be crucial for waste clearance^5,6^ from brain tissue, which lacks a lymphatic system. The glymphatic hypothesis proposes that a brain-wide system connecting the CSF and brain extra-cellular space through perivascular spaces^5^ clears waste, in particular during sleep^6^. The mechanisms behind such clearance are partially unknown^7^ and may be multi-faceted. A variety of theoretical and computational models proposing driving mechanisms for waste include explanations based on peristalsis-driven netflow^8–11^, dispersion^12,13^, multi-compartment diffusion-reaction models^14^, or multiple network poroelasticity ^15,16^. However, the driving mechanisms are still being debated, with no mechanistic model able to explain the variety of clinically and experimentally observed transport phenomena^7,17^.

Few methods today can probe fluid transport in humans. A method for assessing the transport of substances in the CSF and brain is dynamic contrast-enhanced magnetic resonance imaging (MRI): a contrast agent (CA) is injected intrathecally (into the CSF-filled fluid spaces in the spinal canal of the lower back), and the brain is imaged over several days using a combination of multiple MRI sequences^18–29^, see Fig. 1. The CA locally increases the nuclear magnetic relaxation rate^1^ of the sample region by an amount proportional to its concentration; concentration differences can thus be visualized as image contrast using appropriate MR sequences. Intrathecal contrast-enhanced MRI (also called “glymphatic MRI”^22^) is an off-label use of MR contrast agents^30^, relatively time-consuming (≈ 30 min per session for multiple sequences recorded), invasive (CA administration), and typically restricted to patients with underlying CSF pathologies. Although considered safe when using low doses of gadolinium-based contrast medium^25,31,32^, human studies are scarce. MRI data from clinical research projects using glymphatic MRI are not accessible to the public due to privacy concerns and ethical considerations. However, access to image data is crucial for designing data analysis pipelines and three-dimensional, subject-specific computational simulations. These tools support understanding the underlying physics of the complex transport phenomena visualized. For the first time, we can provide such data as a publicly accessible dataset.

**Figure 1.**
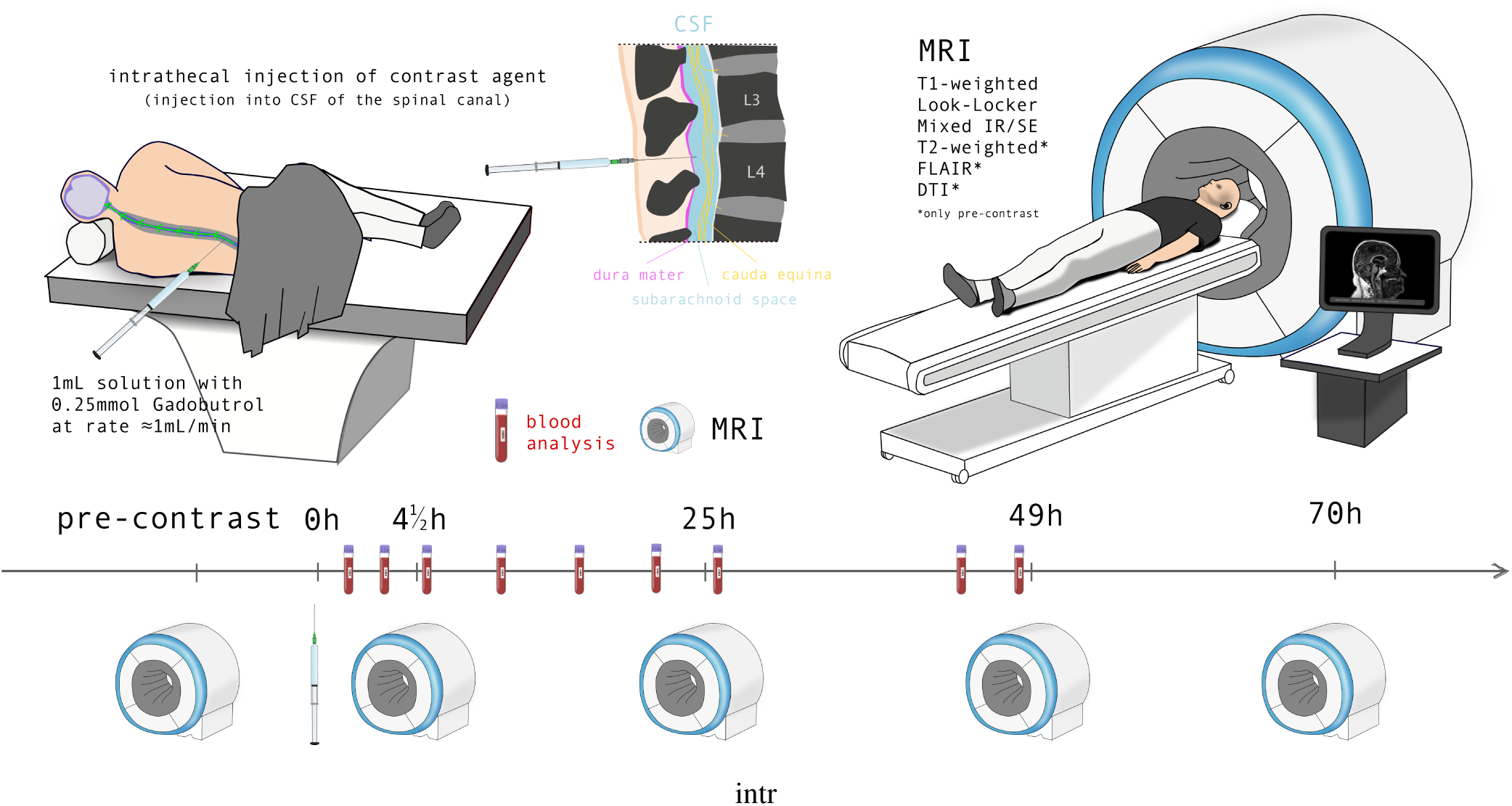
Human brain MRI of CSF tracer evolution over 72h. The dataset contains head images from (contrast-enhanced) magnetic resonance imaging (MRI) with intrathecal injection of contrast agent (CA); the data collection protocol is visualized in the figure. The CA (Gadobutrol) spreads along the spinal canal into the cerebrospinal fluid (CSF) spaces in and around the brain. MR images with different MR sequences (Table 4) are acquired at four subsequent time points as well as before contrast administration (session duration ≈ 30 min and ≈ 1 h pre-contrast). Blood samples are taken between MRI acquisitions and analyzed for CA plasma concentration.

A healthy volunteer participant in a recently started clinical study (see GRIP in Section Methods) on the role of glymphatic clearance in proteinopathies (such as Parkinson’s disease) provided informed consent to conducting a glymphatic MRI study and making the data publicly available. The dataset resulting from the collection procedure shown in Fig. 1 is intended as a data source in data-integrated simulation techniques to research and “reverse-engineer” transport mechanics; it also serves as a starting point to develop data processing frameworks working with glymphatic MRI, or similar data.

An apparent drawback of the presented dataset is that it only contains data from a single individual. Previous studies using glymphatic MRI have shown large inter-individual differences in CSF transport patterns^33^. Hence, the dataset alone does not enable physiological or medical conclusions based on statistics. The dataset is, however, valuable in another way: while data from a single subject is insufficient to construct purely data-driven models, physics-based computational and mathematical modelers can still effectively use the dataset for validation purposes and testing of mechanistic hypothesis; physics-based models emulate the actual transport process in between sparse data^2^. Moreover, a dataset with a single subject also allows for a controlled and repeatable environment for patient-specific method development based on clinical non-synthetic data.

In summary, the purpose of the present dataset is to provide data and simulation scientists with a comprehensive, openly available contrast-enhanced MRI dataset, enabling testing and validation of computational models of solute transport in the brain. Since for the development of mathematical models, quantitative measurements are crucial—even if sparse—we provide blood sample measurements acquired between MRI sessions, cf. Fig. 1 supplementing the raw MRI data. Finally, various post-processing tools and post-processed data that allow the direct use of the data in mesh-based computational simulations are presented.

## Methods

### Inclusion and clinical procedure, MRI data acquisition

A healthy male volunteer in their 60s was recruited for study. He underwent a thorough neurological examination and cognitive tests, which were normal. An MRI examination revealed no more than non-specific white matter hyper-intensities and a tendency towards iron deposition in the basal ganglia. Dementia markers in cerebrospinal fluid were within the normal range (amyloid beta, total tau, and pTau181). Using X-ray guidance, an interventional radiologist performed a lumbar puncture in the lateral position with a thin (G25) atraumatic needle. Proper needle placement was confirmed through the passive release of cerebrospinal fluid, after which an MRI contrast agent (0.25 mmol of gadobutrol, 1 mL) was administered intrathecally. MRI acquisitions were carried out 4.5 h, 1 d (25 h), 2 d (49 h), 3 d (70 h) after contrast agent administration, in addition to pre-contrast MRI.

The study obtained approval from the Regional Ethics Committee (REC #282297) and the Hospital Authority (Data Protection approval #21/19051). The healthy volunteer provided written and oral informed consent, both for participating in the study and for the open-access publication of the full series of MRI scans as part of this publication.

### Blood plasma concentration samples

Intravenous blood samples were obtained at nine different timepoints, and quantification of gadolinium (Gd) in the blood samples were performed by the climate and environmental research institute NILU (Kjeller, Norway), using inductively coupled plasma mass spectormetry, as previously described in^34^. The concentration measurement data points are provided in milligram gadolinium per kilogram plasma. For convenience, we converted the values to molar concentrations of gadobutrol using a plasma density of 1025 kgm^−3^, 157.3 × 10^−3^ kgmol^−1^, and the fact that each gadobutrol molecule contains one gadolinium atom. The results are presented in Table 3. The raw and converted data is included in the dataset (see Data Records).

### MRI data acquisition

The MRI scans acquired include *T*_1_-weighted images, a Look-Locker sequence, and a T2-weighted mixed inversion recovery, spin-echo sequence (Mixed) sequence in all sessions, and *T*_2_-weighted images, FLAIR, and diffusion tensor imaging (DTI) in the pre-contrast session. Scanning was performed on a 3 T Philips Ingenia MRI scanner (Philips Medical Systems, Best, The Netherlands) with a 32-channel head coil. The MRI Sequence parameters are summarized in Table 4. Pre-contrast session sample images are shown in Fig. 2.

**Figure 2.**
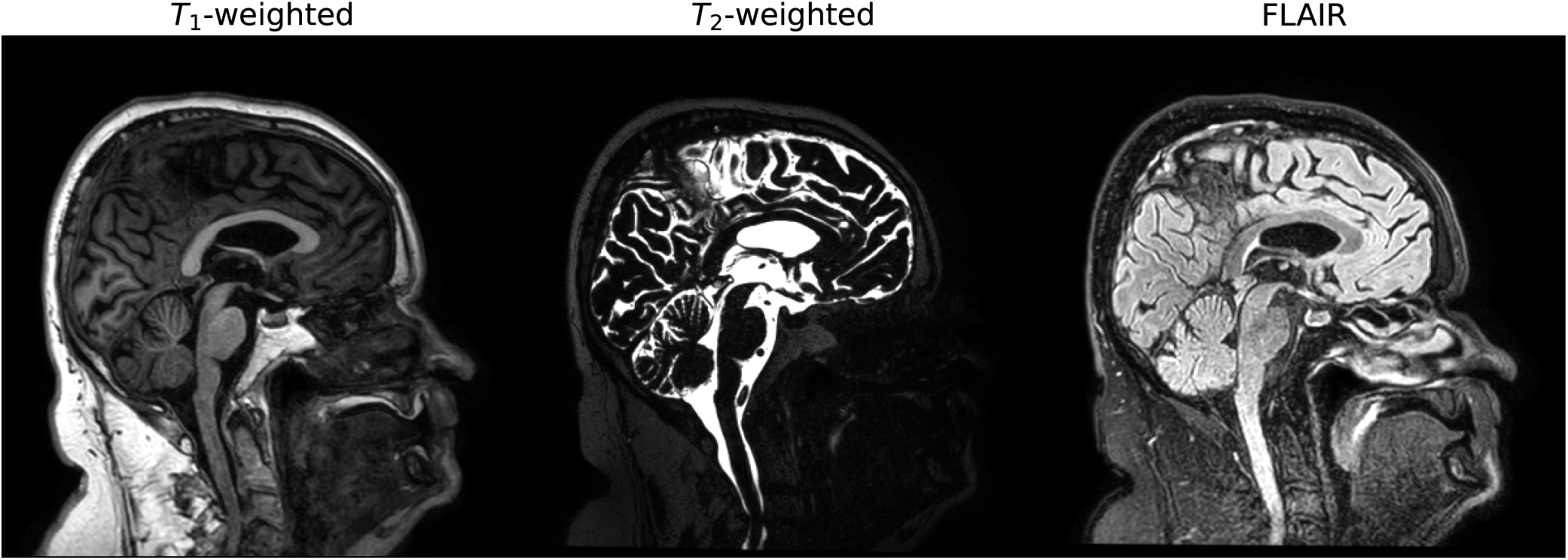
Pre-constrast MRI data. Pre-contrast *T*_1_-weighted, *T*_2_-weighted, and FLAIR images are used for segmentation and cortical reconstruction: the *T*_1_-weighted and FLAIR images by FreeSurfer’s recon-all; the *T*_2_-weighted image to create a CSF mask.

### MRI file format

The image data in this dataset are provided in the NIfTI 1 format. The NIfTI files are obtained by converting the scanner-native Philips-enhanced DICOM format files using the software dcm2niix^3^. Associated with each NIfTI file is a Brain Image

Data Structure (BIDS) file containing metadata in JSON format. Images from the Mixed sequence were converted from Philips-enhanced DICOM to NIfTI using a custom script (see Section Data Records). The script was verified by ensuring it outputs the same floating point values and affine map as dcm2niix for a *T*_1_-weighted image.

### Estimating *T*_1_ times from Look-Locker sequence data

*T*_1_ maps for brain tissue are generated voxel-wise by fitting a curve to the longitudinal magnetization recovery signal from a Look-Locker image sequence^36^ as shown in Fig. 3a for the pre-contrast session. The tissue’s net magnetic moment is flipped anti-parallel to the MRI’s main magnetic field. Following the inversion of magnetic moments, the longitudinal magnetization is expected to follow a curve

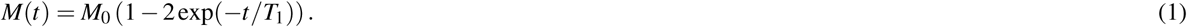

**Figure 3.**
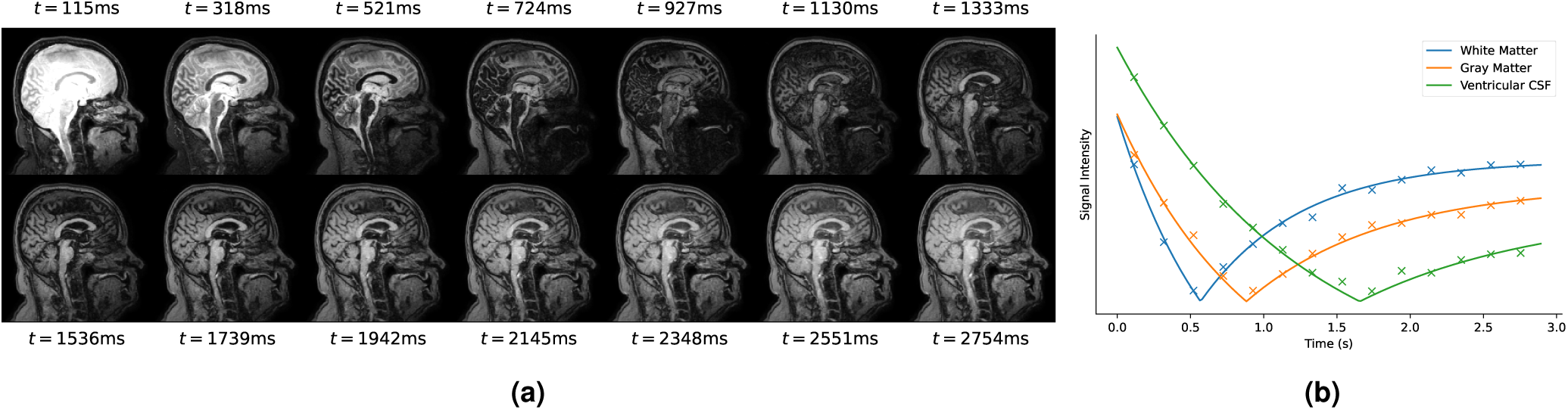
Look-Locker *T*_1_ estimation. **(a)** Sequence of Look-Locker images, representing the magnitude of the longitudinal magnetization for each voxel following an inverting pulse, which flips the net magnetic moment of the tissue to point opposite of the main magnetic field. **(b)** The evolution of the Look-Locker sequence’s signal intensity in three example voxels from different regions (shown as markers), together with the fitted curves on the form 2. For each measurement, 14 data points are acquired over 2750 ms.

However, due to imperfect inversion of the magnetic field, measurement errors, and disturbances induced by signal generation, the *T*_1_ times are typically found by fitting a curve^37^

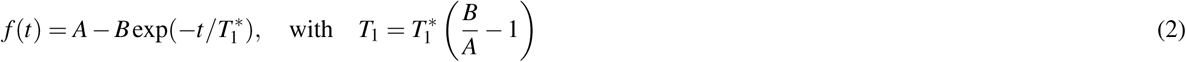

to the measured signal intensities for the generic parameters *A, B* and where 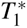 denotes the *apparent T*_1_ time^37^. Instead of directly fitting the above curve to the signal intensities, we reparametrize it to include apriori knowledge about the expected shape and form of the curve,

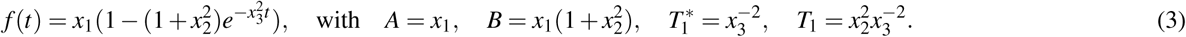

This parametrization guarantees a sign flip for *t* ≥ 0 and a positive *T*_1_ time. The signal intensity time series 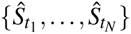 (here *N* = 14) in the Look-Locker sequence images included in this dataset represent the magnitude of the complex longitudinal magnetization value; they are thus positive, cf. Fig. 3b. Therefore, we fit | *f* (*t*)| to the 14 data points per voxel, which we normalize by their local (voxel-wise) maximum value over time to ensure similar order of magnitude. We use scipy.optimize.curve_fit from the SciPy-library^38^ using the Levenberg-Marquardt algorithm. To avoid outliers, we additionally treated extreme values. Voxel values were voided (set to NaN) either before or during the relaxation analysis if either 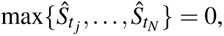 the optimization algorithm does not converge within less than 1000 function evaluations, or if the optimization algorithm raises an exception. After *T*_1_ estimation, we voided voxels with estimated *T*_1_ times outside of the range [50, 5000] ms. Finally, any voided voxels within the head were interpolated from the surroundings. A sagittal view of the resulting *T*_1_ map is shown in Fig. 4.

**Figure 4.**
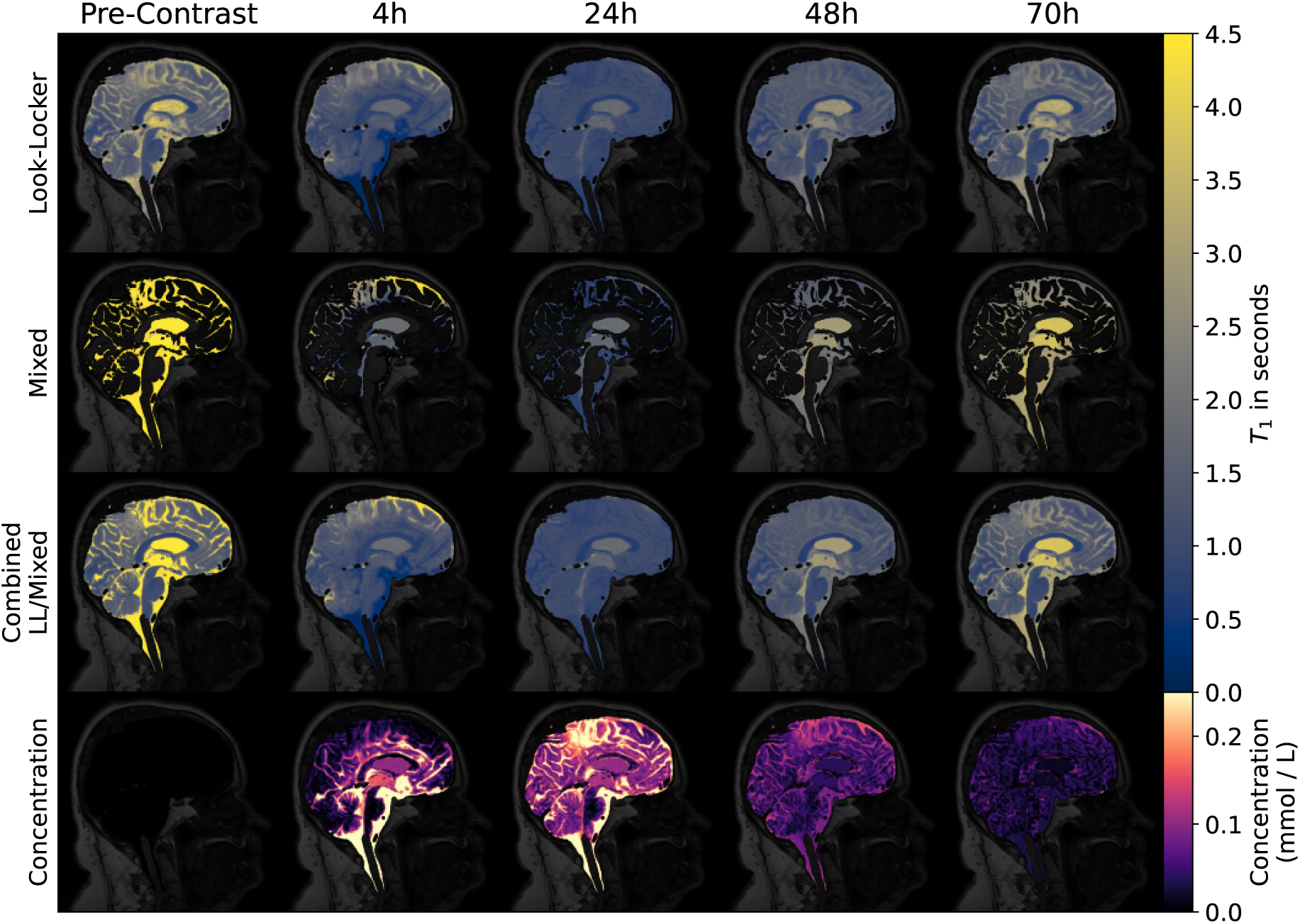
*T*_1_ and concentration maps. *T*_1_ maps estimated from the Look-Locker sequence after outlier removal and interpolation of missing values within the head mask. *T*_1_ maps, i.e., the field of *T*_1_ times (in ms) in cerebrospinal fluid as estimated from the Mixed MRI sequence. Estimated concentrations in subarachnoid space, ventricles, and parenchyma, based on hybrid (Look-Locker / Mixed) *T*_1_ maps for all five sessions.

### Estimating *T*_1_ times from the Mixed sequence

The Lock-Locker estimation yields poor results (see Section Technical Validation) if the data acquisition period is much shorter than the *T*_1_ relaxation time of the sample (see, for instance, the green curve (ventricular CSF) in Fig. 3(b), which at the end of the acquisition period is still far from equilibrium magnetization). Longer acquisition periods are technically possible but lead to longer MRI sessions (the Lock-Locker sequence with a data acquisition time of 2750 ms takes ≈ 12 min, cf. Table 4) possibly causing patient discomfort. The mixed inversion recovery and spin echo sequence (Mixed), introduced in^39^, is designed to enable the estimation of long *T*_1_ times (e.g., CSF) more accurately and faster than with the Look-Locker sequence. Two images are acquired: a spin echo signal with signal *S_SE_* and an image inversion recovery image with signal *S*_IR_. The signals are estimated by

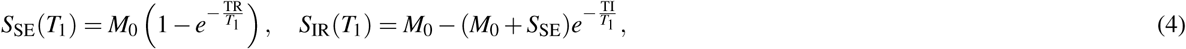

where *M*_0_ is the equilibrium magnetization of the tissue, TI is the inversion time and TR is the repetition time^40^. To estimate *T*_1_ maps from these two signals, one observes that the ratio

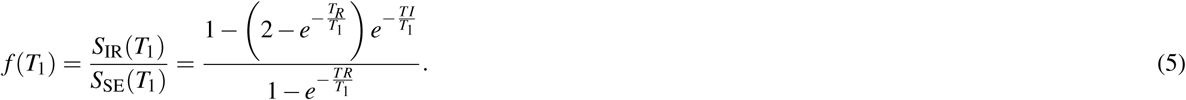

is a monotonically decreasing function and therefore invertible: by computing the ratio *f* (*T*_1_) from measured data, we can get *T*_1_ by solving the nonlinear Eq. (5). The *T*_1_ times estimated from the Mixed sequence data are accurate for high *T*_1_ values (CSF, low tracer concentration) but become increasingly inaccurate for low *T*_1_ (gray matter or high tracer concentration); see Section Technical Validation. To only retain *T*_1_ estimates for fluid-filled spaces with low contrast, a mask was generated by applying Yen’s thresholding algorithm^41^ to the spin echo images. *T*_1_ values in voxels outside this mask were voided (set to NaN). Figure 4 shows the estimated *T*_1_ maps after masking for each session.

### Registration

All images were registered and resampled to the image space of the pre-contrast *T*_1_-weighted image using the software greedy^42^ (https://github.com/pyushkevich/greedy) for rigid registration of images. Rigid registration of an input image to a target image results in a 4 × 4 matrix representing an affine transformation in homogeneous form for reuse. We used the default parameters of the registration software except for the following MRI-sequence-specific modifications: The pre-contrast *T*_2_-weighted images were registered directly to the target space using a normalized mutual information (NMI) loss function. Each of the post-contrast *T*_1_-weighted images was registered directly to the pre-contrast *T*_1_-weighted image, using a normalized cross-correlation (NCC) loss function with a 5 × 5 × 5 neighborhood. The Look-Locker *T*_1_ maps were resampled to the target space by first registering the corresponding inverted *T*_1_ map images to the target space using an NCC 5 × 5 × 5 loss function and then applying the output transformation to the *T*_1_ maps. The *T*_1_ maps estimated from the Mixed sequence were resampled to the target space by first registering the mixed spin-echo volume to the *T*_2_-weighted image using an NCC loss function with a 5 × 5 × 5 neighborhood and applying the output transformation to the *T*_1_ maps. Following tensor reconstruction, the mean diffusivity was registered to the pre-contrast *T*_1_-weighted image. Finally, the DTI data was resampled to the target space by first registering the estimated mean diffusivity image to the already registered *T*_2_-weighted image, with the NMI loss function. The estimated transform from the preceding step was then used to resample the mean-diffusivity, fractional anisotropy, eigenvalues, and eigenvectors into the image space of the pre-contrast *T*_1_-weighted image. Since greedy only works with 3D MRI data, the eigenvectors are split into components, resampled component-wise, merged to a 4D structure, and renormalized to unit vectors. The resampled eigenvectors and eigenvalues were then used to reconstruct the symmetric diffusion tensors in the target space, with all nine components stored row-wise.

### Hybrid *T*_1_ maps

The *T*_1_ times estimated from the Mixed sequence are expected to be most accurate for high *T*_1_ values (e.g., CSF-filled spaces such as the subarachnoid space and the ventricles), whereas the Look-Locker estimates are suitable for small *T*_1_ (e.g., brain tissue and high tracer concentrations). To distinguish CSF from tissue, we created a CSF mask *M_CSF_*. The mask was created using Yen’s thresholding method^41^ (binary segmentation) on the pre-contrast *T*_2_-weighted image (Fig. 2 middle pane) registered and resampled into the reference image space. After binary segmentation, we discarded all but the largest connected region. While the Mixed sequence is designed for large *T*_1_ times of pre-contrast CSF (*>* 4 s), the contrast agent reduces *T*_1_ significantly. Post-contrast *T*_1_ times can reach values well below 1 s in CSF regions with high contrast agent concentrations. Based on the analysis described in Section Technical Validation, we choose a threshold of *T*_1_ = 1500 ms to create the hybrid *T*_1_ map

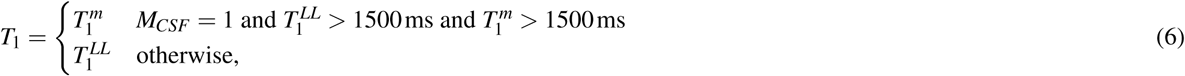

where 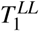 and 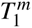 denote the registered *T*_1_ maps estimated from Look-Locker and the Mixed sequence, respectively. Figure 4(third row) shows the hybrid *T*_1_ map for the pre-contrast session. The hybrid map is used to estimate tracer concentrations.

### Concentration estimation

Tracer concentrations *C* are estimated voxel-wise from the hybrid *T*_1_ maps for each session, based on the relation

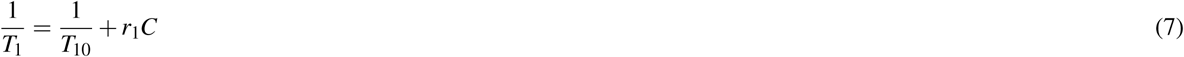

where *T*_10_ is the native *T*_1_ time of a given voxel, and *r*_1_ = 3.2 s^−1^ Lmmol^−1^ is the *T*_1_ relaxivity, estimated in^43^. Since we are only interested in intracranial concentrations, we mask the voxels outside the cranium: starting with the binary segmentations of the CSF and the brain, denoted *M_CSF_* and *M_B_*, respectively, we create an initial background mask by inverting the union of CSF and brain segmentations. Next, the largest island is extracted from the background mask to remove potential gaps between the CSF and brain segmentations. Finally, we use a binary opening algorithm with a ball of radius 3 as the structuring element. The concentrations estimated from the hybrid Look-Locker/Mixed *T*_1_ maps are shown for each session in Fig. 4.

### Diffusion Tensor Images

The dataset contains dynamic diffusion tensor images (15 directions, *b* = 0 and 1000 smm^−2^, AP) from 10 different time points. The images were initially corrected for susceptibility distortions and eddy-currents using FSL’s^44^ topup^45,46^, and eddy^47^. After initial correction, the eigenvalues and eigenvectors of the diffusion tensors were estimated using dtifit. Figure 5 shows the mean-diffusivity, fractional anisotropy, and color-coded fractional anisotropy after registration. Finally, the diffusion image was checked for invalid diffusion tensors based on whether any eigenvalue is negative or whether the fractional anisotropy lies outside the valid range [0, 1]; invalid tensors were replaced by the nearest valid diffusion tensor. We refer to [48, Chapter 5] for further details on the post-processing steps.

**Figure 5.**
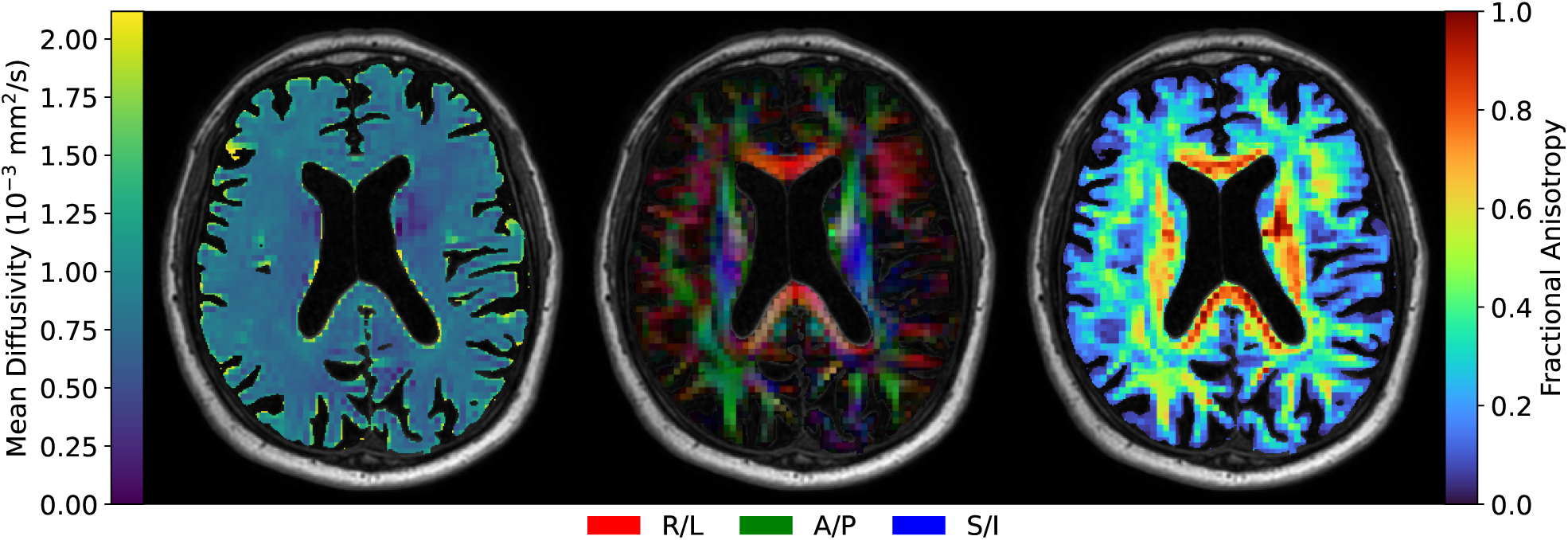
Diffusion tensor imaging. DTI data represented by (left) mean diffusivity, (right) fractional anisotropy and (middle) directional color-coded fractional anisotropy, with RGB-channels *FA* × (|*V*_1*x*_|, |*V*_1*y*_||*V*_1*z*_|) for the normalized principal eigenvector *V*_1_. Red goes in the (R)ight-(L)eft direction, green along the (A)nterior-(P)osterior and blue along the (S)uperior-(I)nferior axis.

### Normalization of *T*_1_-weighted images

Normalized, post-contrast, *T*_1_-weighted images have previously been used to investigate tracer enrichment in brain tissue following intrathecal injection of contrast agent in a range of studies^22,23,27,33,49^. Following the strategy in^50^, the images were normalized by scaling the signals relative to the median signal in a reference region in the (left) orbital eye fat. To automatically generate the reference region, we identified three regions (1012: ctx-lh-lateralorbitofrontal; 1027: ctx-lh-rostralmiddlefrontal; 1033: ctx-lh-temporalpole) from the FreeSurfer segmentation aparc+aseg.mgz, one for each coordinate axis and manually align them with the reference regions in the direction of their respective coordinate axes. From these regions, a point in the vicinity of the left orbital fat was found by intersecting the three planes, which lie perpendicular to the coordinate axes and pass through the corresponding FreeSurfer region. The intersecting point was used as the center of a multivariate Gaussian probability distribution with a diagonal covariance matrix chosen such that most of the orbital eye fat is assigned a high probability density, as shown in Fig. 7a. The distribution was multiplied with the signal intensities of the *T*_1_-weighted image, leaving a focused view of the original image, as shown on the left of Fig. 7b. Thereafter, a first estimate of the reference region was generated by using Yen’s thresholding algorithm^41^ implemented in skimage.morphology. As a post-processing step, we applied binary erosion to the binary segmentation and removed all but the largest island. This procedure was performed for the *T*_1_-weighted image from each session (after registration to the pre-contrast image), and the final reference region was taken as the intersection between the reference regions generated from each image.

### Cortical reconstruction and segmentation

FreeSurfer’s^51^ cortical reconstruction pipeline recon-all (FreeSurfer version 7.4.1) was used to create surfaces for generating the finite element mesh and segmenting the different regions of the subject’s brain. The FreeSurfer-based pipeline was run for the pre-contrast session, taking the *T*_1_-weighted image as main input (as shown in the leftmost image in Fig. 6, together with the FLAIR image which helps for the pial surface reconstruction. Brain segmentation has two main purposes: First, it enables us to compute region-specific quantities, such as total tracer mass within a specific region, allowing for quantitative analysis of tracer movements. Second, segmentation forms the basis for boolean arrays, which allow us to process brain tissue differently from CSF-filled spaces. We also include the output of the FastSurfer pipeline, a fully compatible FreeSurfer alternative, using deep learning for segmentation and a spectral projection algorithm for surface reconstruction^52^.

**Figure 6.**
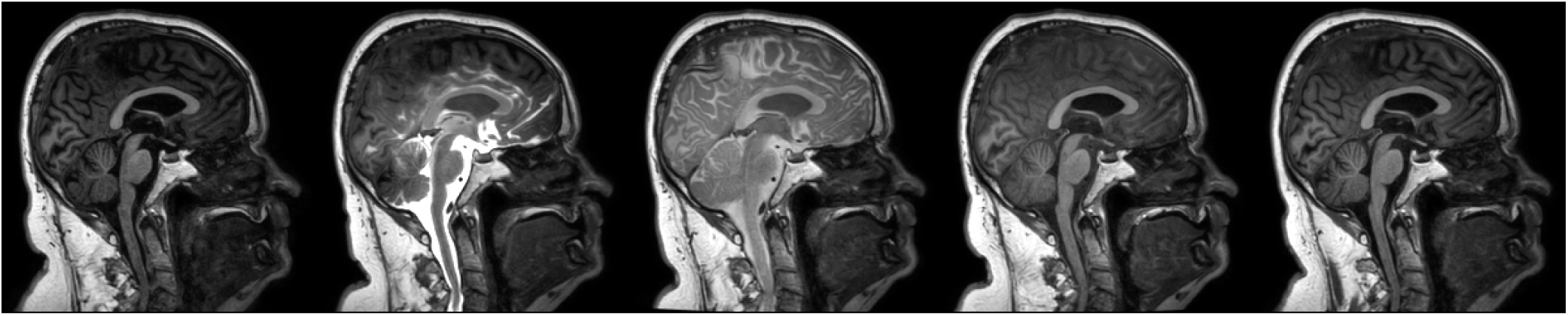
*T*_1_-weighted images. *T*_1_-weighted images before normalization, axial view, pre-contrast and post-contrast injection at times 4 h, 24 h, 48 h and 70 h, ordered chronologically from left to right.

**Figure 7.**
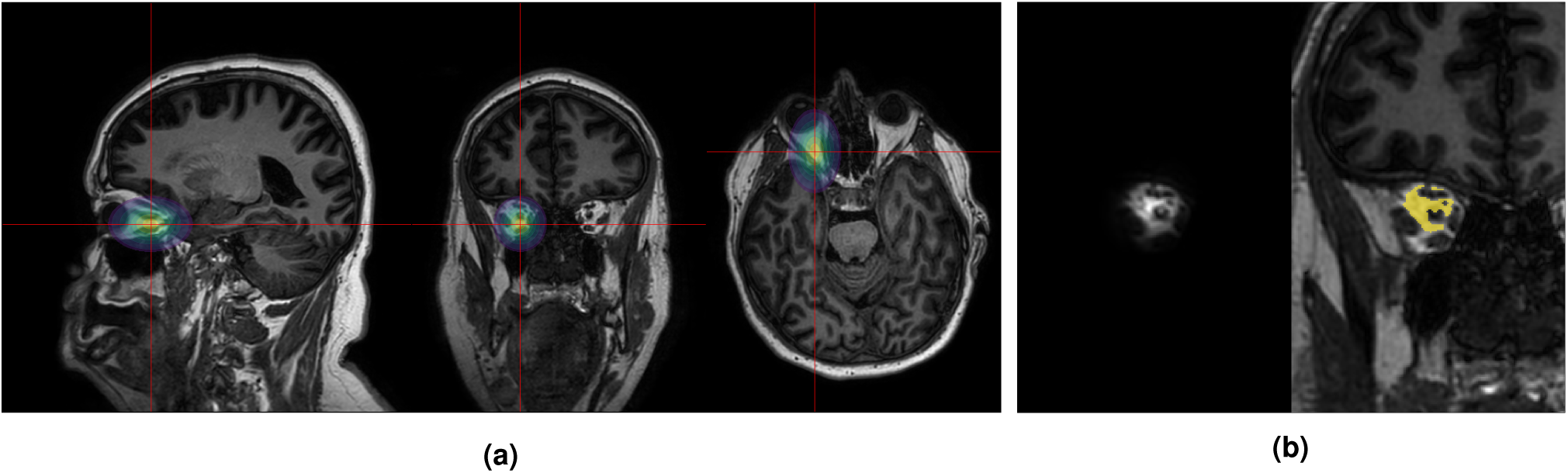
Normalization of *T*_1_-weighted images. An illustration of the automated mask generation routine for the reference region: **(a)** The red lines represent planes along each coordinate axis. They intersect at a point close to the left orbital fat. The contours surrounding the points represent a Gaussian distribution centered at the intersecting point. **(b)** The left part shows the product between the Gaussian distribution from (a) and the *T*_1_-weighted image, leaving an image consisting mainly of the orbital fat. A mask is generated from the corresponding image for each session. The masks are intersected to create the reference region, shown in yellow on the right image.

### Mesh generation

A 3D computational mesh (tetrahedral volume mesh) of the cerebrum, as shown in Fig. 8a, was generated using a process based on the one described in [48, Ch. 4]. The process relies on surface meshes of the ventricles, subcortical gray matter, the pial surface, and surfaces for the interface between the gray and white matter. The pial and white matter surfaces (for each hemisphere) were created by the FreeSurfer recon-all pipeline. The ventricles (as shown in Fig. 8c) and subcortical gray matter surfaces were extracted as contour surfaces based on the FreeSurfer segmentation aseg.mgz. After meshing, the ventricles were removed, so only the brain tissue remains. The subcortical gray matter structures can be seen in the subdomain labels of Fig. 8b. The pre-processing of surfaces and the mesh generation uses the Python libraries pyvista^53^ and SVMTK^4^. Further details on the procedure may be found in the script src/brainmeshing/mesh_generation.py in the repository https://github.com/jorgenriseth/gMRI2FEM (also see Section Code Availability).

**Figure 8.**
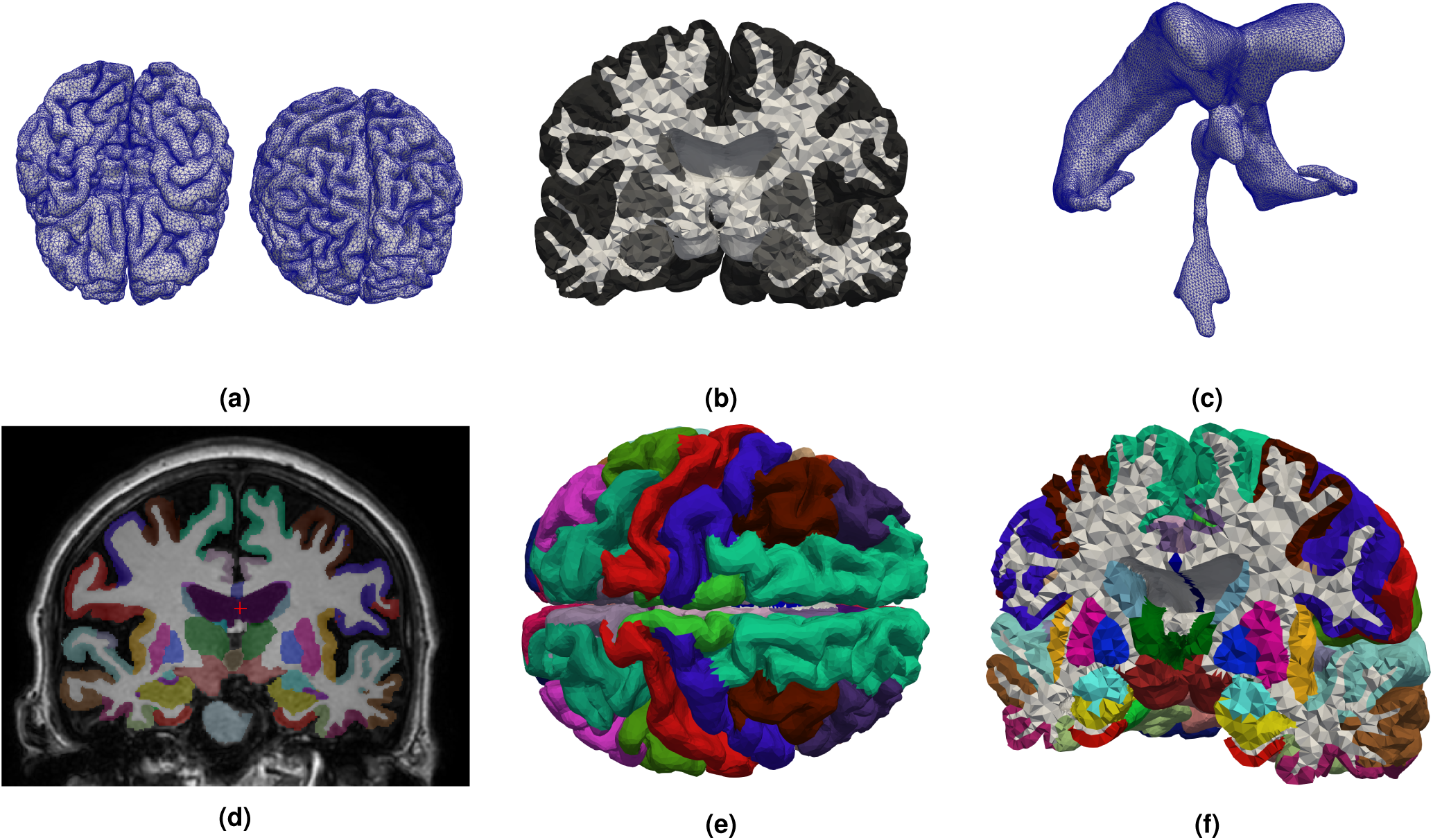
Meshes and subdomain tags. **(a)** top and bottom view of the brain mesh with edges to highlight each of the cells. **(b)** cell tags defining subdomains based on the enclosing surfaces during mesh generation. Cortical gray matter is dark gray, sub-cortical gray is light gray, and white matter is white. **(c)** ventricle surface mesh. **(d-f)** subdomain tags created by mapping the FreeSurfer MRI segmentation aparc+aseg in different views.

### Mapping FreeSurfer segmentation data onto the mesh

The mesh can be subdivided into subdomains corresponding to the FreeSurfer segmentation data such as shown in Figs. 8d to 8f for the aseg+aparc-segmentation. Each cell is labeled according to the most common label in the voxels in a neighborhood around the cell midpoint, with an additional check to ensure that each label is contained within only one of the subdomains illustrated in Figs. 8d to 8f originating from the meshing procedure. We refer to [48, chapter 4] for further details.

### Mapping function data onto the mesh

Mesh vertices are associated with two types of concentration data: one representing the concentration field within the CSF at the brain surface and one representing the tissue concentrations within the brain. Both can be represented as piecewise-linear, continuous finite element functions across the entire domain (setting inner nodes to zero for the surface data).

The surface concentration data is intended to be used to derive boundary conditions for mesh-based transport simulations. The degrees of freedom of the basis functions that are associated with the boundary vertices are assigned the median concentration of the ten nearest voxels within the CSF, as defined by the previously described CSF mask. All internal degrees of freedom are assigned 0. The surface concentration for all sessions is shown in Fig. 9. The internal concentrations *u* are mapped from the MRI-image *C* by an approximate Galerkin projection onto the space of continuous, piecewise linear functions *V_h_* on the mesh Ω*_h_*. The Galerkin projection is done by solving the variational problem

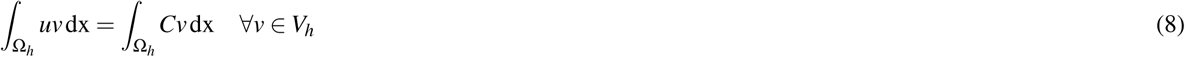

where we interpret *C* as a piecewise constant function on a quadrilateral mesh containing the brain mesh Ω*_h_*. The assembly of the algebraic system corresponding to the variational equation (8) entails using a numerical quadrature rule for each cell in the mesh. We approximate the right hand side of the variational form using Gaussian quadrature with quadrature degree 6.

**Figure 9.**
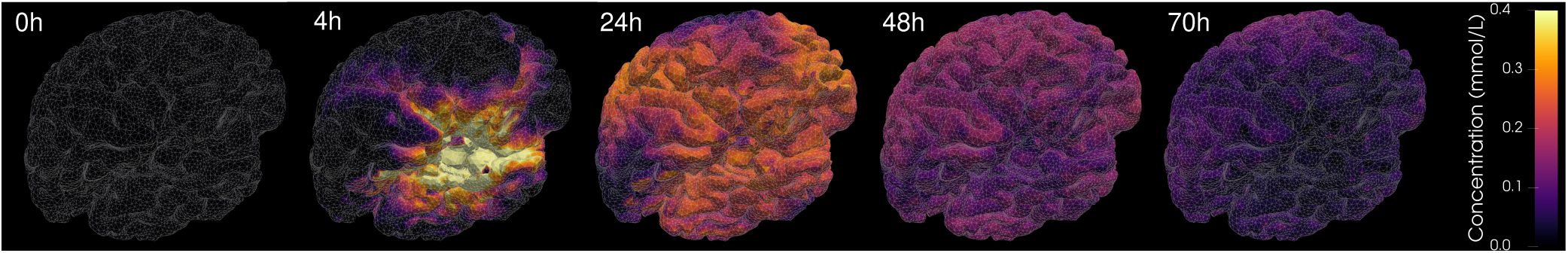
CSF tracer concentration mapped onto brain surface. Concentration shown in the range 0.0 mmol L^−1^ to 0.4 mmol L^−1^ as given by a piece-wise linear function mapped onto the surface of the brain volume mesh. Note that the data also contains data outside the range. These outliers are not shown in this figure.

In addition to the concentrations, data from DTI are also included with the mesh. The data includes mean diffusivity, fractional anisotropy, and diffusion tensors. However, these quantities are represented as cell fields, and the values are assigned from the neighborhood of the cell midpoint. The mean diffusivity and fractional anisotropy are assigned the median value of the ten nearest neighbors, whereas the tensors are assigned from the single nearest voxel.

## Data Records

All records are available as a Zenodo-archive at https://zenodo.org/records/14266867. The files are split into five different zip-archives, as further detailed below. All files with extension *.nii.gz* represent MRI-images in NIfTI-1 file format.

**mri-dataset.zip** Contains the MRI images output by dcm2niix with meta-data.

- **mri_dataset/**

**–** timetable.tsv - Timetable of MRI sequences given in seconds relative to time of contrast injection.
**–** blood_concentrations.csv - Gadolinium concentrations measurements from blood plasma samples.
- **mri_dataset/sub-01/ses-01/anat/** - Contains anatomical MRI available only for the pre-contrast session.

**–** sub-01_ses-01_FLAIR{.nii.gz,.json} - FLAIR MRI with BIDS sidecar.
**–** sub-01_ses-01_T2w{.nii.gz,.json} - T2-weighted MRI with BIDS sidecar.
- **mri_dataset/sub-01/ses-XX/anat/** - Contains anatomical MRI available only for all 5 sessions.

**–** sub-01_ses-XX_T1w{.nii.gz,.json} - T1-weighted MRI with BIDS sidecar.
**–** sub-01_ses-XX_acq-looklocker_IRT1{.nii.gz,.json} - Look-Locker sequence (4D MRI) with sidecar.
**–** sub-01_ses-XX_acq-looklocker_IRT1_trigger_times.txt - Text file with the trigger times in milliseconds for the corresponding Look-Locker sequence.
- **mri_dataset/sub-01/ses-XX/mixed/** - Contains Mixed data available for all 5 sessions

**–** sub-01_ses-XX_acq-mixed.json - BIDS sidecar accompanying all mixed MRI data.
**–** sub-01_ses-XX_acq-mixed_IR-corrected-real.nii.gz - Mixed inversion recovery MRI.
**–** sub-01_ses-XX_acq-mixed_SE-modulus.nii.gz - Mixed spin-echo MRI.
**–** sub-01_ses-XX_acq-mixed_T1map_scanner.nii.gz - T1-map generated by scanner from mixed sequence.
**–** sub-01_ses-XX_acq-mixed_meta.json - Additional metadata needed for T1-map generation from Mixed.
- **mri_dataset/sub-01/ses-01/dwi/** - Contains diffusion-weighted MRI data available only for the pre-contrast session.

**–** sub-01_ses-01_acq-multiband_sense_dir-AP_DTI{.nii.gz,.json,.bval,.bvec} - Diffusion weighted measurements with BIDS sidecar, and bval/bvec-files describing strength and direction of the gradient field.
**–** sub-01_ses-01_acq-multiband_sense_dir-AP_DTI_ADC.nii.gz - Apparent diffusion coefficient output by dcm2niix.
**–** sub-01_ses-01_acq-multiband_sense_dir-PA_b0{.nii.gz,.json} - Reference MRI with BIDS sidecar, taken with opposing phase-encoding direction of the DTI-data for correcting susceptibility induced distortions.

**mri-dataset-precontrast-only.zip** Contains the same data as **mri_dataset.zip**, but only for the pre-contrast session (ses-01).

**mri-processed.zip** Contains MRI data derived from the raw data.

- **mri_dataset/derivatives/sub-01/ses-01/dwi/** - Contains DTI-data generated by fitting diffusion tensors using dtifit.

**–** sub-01_ses-01_dDTI_FA.nii.gz - Fractional anisotropy of the diffusion tensors.
**–** sub-01_ses-01_dDTI_L{1,2,3}.nii.gz - Eigenvalues of the diffusion tensors.
**–** sub-01_ses-01_dDTI_MD.nii.gz - Mean diffusivity of the diffusion tensors.
**–** sub-01_ses-01_dDTI_V{1,2,3}.nii.gz - Eigenvectors of the diffusion tensors.
**–** sub-01_ses-01_dDTI_tensor.nii.gz - 6-component (symmetric) diffusion tensors, ordered (*D*_*xx*, *D*_*xy*, *D*_*xz*, *D*_*yy*, *D*_*yz*, *D*_*zz*).
- **mri_dataset/derivatives/sub-01/ses-XX/** - Contains DTI-data derived directly from the raw images.

**–** sub-01_ses-XX_acq-looklocker_T1map.nii.gz - T1-map estimated from Look-Locker.
**–** sub-01_ses-XX_acq-looklocker_T1map_nICE.nii.gz - T1-map estimated from Look-Locker using nordicICE.
**–** sub-01_ses-XX_acq-mixed_T1map.nii.gz - T1-map estimated from the mixed-sequence.
- **mri_processed_data/sub-01/T1maps/** - Contains combined Look-Locker, Mixed T1-maps.

**–** sub-01_ses-XX_T1map_hybrid.nii.gz - Hybrid T1-map combining T1maps from Look-Locker and Mixed.
- **mri_processed_data/sub-01/T1w_normalized/** - Contains normalized T1-weighted images.

**–** sub-01_ses-XX_T1w_normalized.nii.gz - T1-weighted MRI after normalization relative to orbital fat signal.
- **mri_processed_data/sub-01/concentrations/** - Contains concentation data.

**–** sub-01_ses-XX_concentration.nii.gz - MRI representing intracranial tracer concentrations.
- **mri_processed_data/sub-01/dwi/** - Contains DTI-data processed following registration.

**–** sub-01_ses-01_dDTI_cleaned.nii.gz - 6-component diffusion tensor after replacement of invalid tensors.
- **mri_processed_data/sub-01/registered/** - Contains MRI data registered to the image space of the reference the pre-contrast *T*_1_-weighted image. All MRI images within mri_processed_data/sub-01 go via this directory.

**–** sub-01_ses-XX_sequence_registered.nii.gz - Represent any MRI registered and resliced into the reference space.
- **mri_processed_data/sub-01/segmentations/** - Contains various segmentations of the brain and CSF spaces.

**–** sub-01_seg-aparc+aseg_refined.nii.gz - FreeSurfer segmentation aparc+aseg resampled into reference image space.
**–** sub-01_seg-aseg_refined.nii.gz - FreeSurfer segmentation aseg resampled into reference image space.
**–** sub-01_seg-csf-aparc+aseg.nii.gz - Segmentation of CSF-space based on nearest neighouring label in aparc+aseg.
**–** sub-01_seg-csf-aseg.nii.gz - Segmentation of CSF-space based on nearest neighouring label in aparc+aseg.
**–** sub-01_seg-csf-wmparc.nii.gz - Segmentation of CSF-space based on nearest neighouring label in wmparc.
**–** sub-01_seg-csf_binary.nii.gz - CSF-mask in reference space.
**–** sub-01_seg-intracranial_binary.nii.gz - Intracranial mask generated from combining CSF-mask and binary representation of aseg.
**–** sub-01_seg-refroi-left-orbital_binary.nii.gz - Left orbital fat reference region mask.
**–** sub-01_seg-wmparc_refined.nii.gz - FreeSurfer segmentation wmparc resampled into reference image space.
- **mri_processed_data/sub-01/transforms/** - Contains affine transformation matrices used to register and reslice MR images into reference space.

**–** sub-01_ses-XX_sequence.mat - Affine transformation matrix for reslicing the given MRI sequence into the reference space.

**freesurfer.zip** FreeSurfer-data.

- **mri_processed_data/freesurfer/sub-01/** - Contains output of the FreeSurfer recon-all-pipeline. See

https://surfer.nmr.mgh.harvard.edu/fswiki/ReconAllOutputFiles for further details.

**surfaces.zip**

- **mri_processed_data/sub-01/modeling/surfaces/** - Contains processed surfaces used as input for the SVMTK meshing algorithm.

**–** lh_pial.stl - Left hemisphere pial surface.
**–** rh_pial.stl - Right hemisphere pial surface.
**–** subcortical_gm.stl - Subcortical gray matter surfaces.
**–** ventricles.stl - Ventricular system surfaces.
**–** white.stl - Merged white-matter surfaces of right and left hemispheres.

**mesh-data.zip**

- **mri_processed_data/sub-01/modeling/resolution32/** - 3D tetrahedral meshes with mapped MRI data.

**–** data.hdf - FEniCS-compatible HDF5 file with mesh and function-representation of MRI data.
**–** data.vtk - VTK Legacy Format file (binary) with 3D mesh and mapped MRI data.
**–** data.vtu - VTK Unstructured Grid XML file (ASCII) with 3D mesh and mapped MRI data.
- **mri_processed_data/sub-01/modeling/resolution32/mesh_xdmfs/** - Contains XDMF files created during the 3D meshing process; for visualization of mesh and subdomain data.

**–** boundaries{.xdmf,.h5} - XDMF3 file with accompanying HDF5 file containing facet-tags generated during meshing process.
**–** mesh{.xdmf,.h5} - XDMF3 file with accompanying HDF5 file containing the 3D mesh.
**–** subdomains{.xdmf,.h5} - XDMF3 file with accompanying HDF5 file showing cell tags generated during meshing process.

We provide post-processing and reuse example code in addition to the main MRI dataset (see Code Availability).

## Technical Validation

### *T*_1_ estimates

To verify the *T*_1_ times estimated from the Look-Locker sequence, we compare our *T*_1_ map with the *T*_1_ map generated by the software nordicICE^5^. Both maps were registered to the pre-contrast *T*_1_-weighted image. For the comparison, we consider only intracranial voxels for which the hybrid *T*_1_ maps rely on the Look-Locker estimates, cf. Eq. (6). The results are shown in Fig. 10. Figure 10a shows a voxel-wise comparison between the estimated *T*_1_ times and the voxel-wise relative error distribution for each session. As part of the Mixed sequence, the proprietary scanner software generates a *T*_1_ map with an upper threshold at 4095 ms and thus misrepresents typical *T*_1_ times of CSF. However, it allows us to verify our *T*_1_ map against another implementation for values below this threshold. A voxel-wise comparison of the estimates is shown for the last session in Fig. 10b, for which most of the *T*_1_ times of the CSF are below the threshold that is clearly visible as a horizontal line in the data. The mean value and standard deviation of the *T*_1_-estimates in gray matter, white and how they compare to previous studies are listed for gray matter, white matter, and CSF in Table 2.

**Figure 10.**
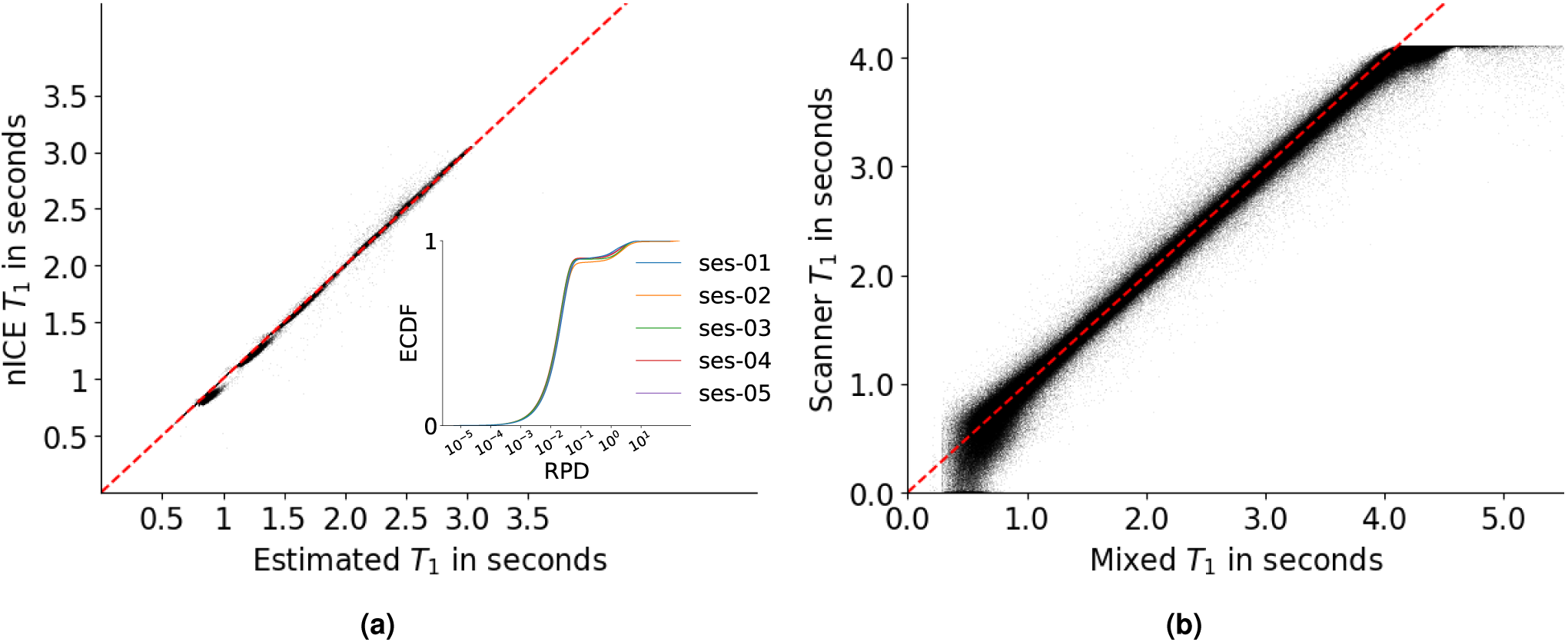
Comparison to different *T*_1_ analysis software. The figure compares our *T*_1_ maps and those created using nICE relaxation analysis, considering only voxels within the intracranial mask. **(a)** Estimated *T*_1_ time vs. the *T*_1_ time of the nICE-analysis from the pre-contrast Look-Locker sequence. **Inset**: Empirical cumulative distribution (ECDF) of the absolute value of the relative percentage difference (RPD) defined as 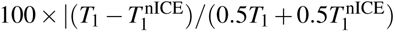 between estimated *T*_1_ times and nICE for each of the sessions (log-scale). **(b)** Estimated *T*_1_ time vs. the scanner-generated *T*_1_ time from the first post-contrast Mixed sequence. The scanner’s intensity cut-off causes the deviation for large *T*_1_ values and is the reason for using our custom algorithm.

To estimate the accuracy of Look-Locker (LL) and Mixed approaches to *T*_1_ time and concentration estimation, we perform an error propagation analysis for the Look-Locker and Mixed post-processing algorithms. From this analysis, we want to motivate the chosen threshold in Eq. (6).

For the Mixed sequence, we model the inversion recovery signal *S*_IR_ and the spin echo signal *S*_SE_ as in Eq. (4). Simulated image intensities 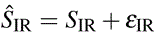 and 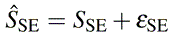 are obtained with TR = 9600 ms, TI = 2650 ms, the error term *ε*_SE_ modeled by a Rayleigh distribution (centered Rice distribution) with standard deviation *σ*_SE_ = max(*S*_SE_)*/*SNR, and *ε*_IR_ modeled by a centered normal distribution with standard deviation *σ*_IR_ = max(*S*_IR_)*/*SNR. For the Look-Locker sequence, the signal over time is modeled by Eq. (3) setting *x*_1_ = 1, *x*_2_ = 2^−1^*^/^*^2^, *x*_3_ = *x*_2_*/T*_1_. To obtain simulated signal intensities, we sample the signal at 14 equidistant time points in *T* = [0 ms, 2600 ms] and add an error term modeled by a Rayleigh distribution with standard deviation max*_t_*_∈*T*_ ( *f* (*t*))*/*SNR where we assume that the errors at different time points are uncorrelated. Monte Carlo type simulations are conducted as follows: (1) For 100 different concentrations (“True”), compute the corresponding *T*_1_ times, and simulate 50 signals (for Look-Locker 50 times 14 data points). (2) Estimate *T*_1_ time and concentration (“Estimated”) from the simulated data described in Section Methods. For the simulations, we mapped concentrations to *T*_1_ time and vice versa with Eq. (7) assuming *T*_10_ = 4500 ms.

Estimated versus true concentrations and *T*_1_ times for SNR = 25 are shown in Fig. 11. Due to the relatively short acquisition time of 2600 ms used for the the Look-Locker sequence, *T*_1_ estimates are increasingly affected by noise with increasing *T*_1_ time. For *T*_1_ *<* 1500 ms and *c >* 0.1 mmol estimates are most accurate. On the other hand, the Mixed sequence is increasingly affected by noise for decreasing *T*_1_ times and increasing concentrations. The concentration estimate appears most accurate for *c <* 0.1 mmol such that the two sequences complement each other.

**Figure 11.**
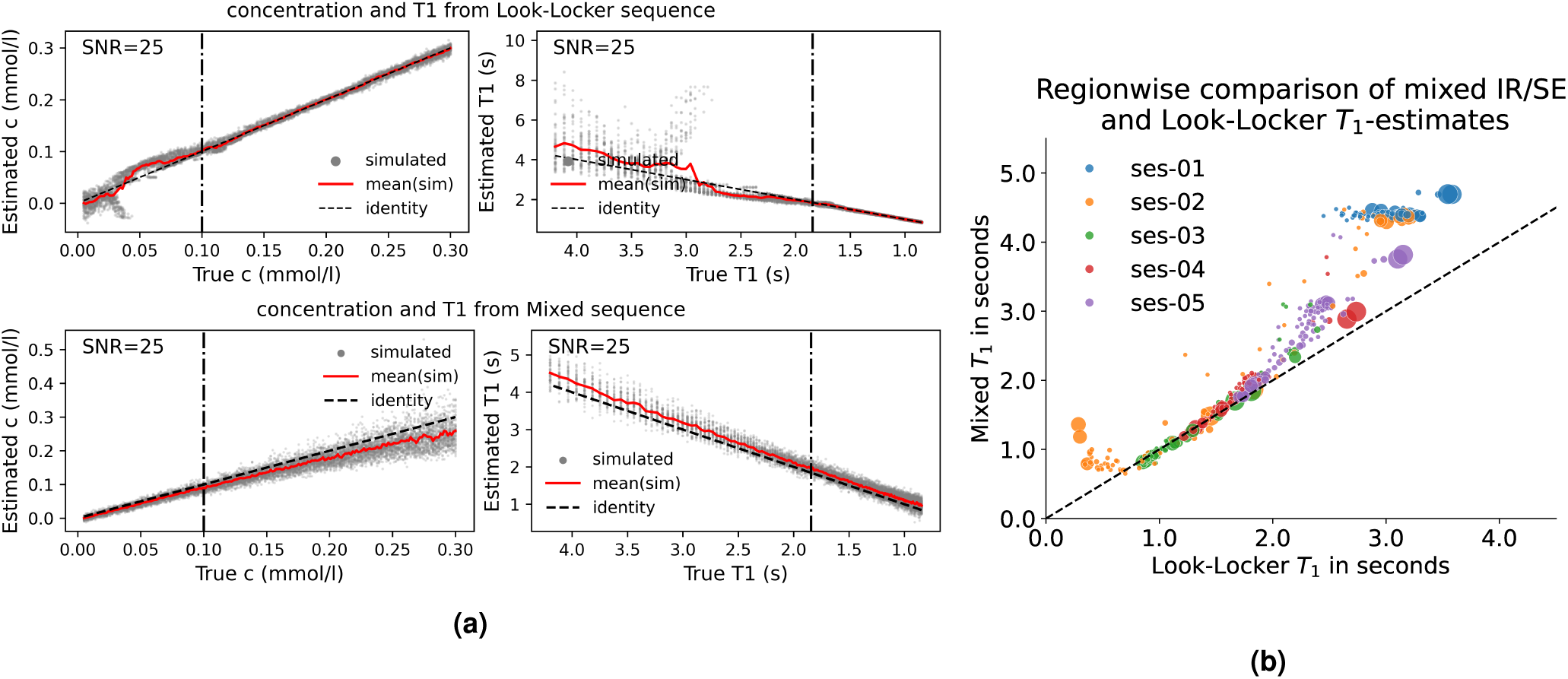
LL/Mixed *T*_1_ estimation: sensitivity to noise. **(a)** Estimated versus true *T*_1_ and concentration values when propagating noisy data through the estimation pipeline. Increasing concentration values correspond to decreasing *T*_1_ times. The relationship is nonlinear, Eq. (7). The dashed line marks a chosen threshold value of the concentration (and corresponding *T*_1_ time) above which concentration estimates with the Look-Locker sequence appear more accurate than estimates with the Mixed sequence (and vice versa below the threshold). The figure is obtained with the script noise/plot_noise_combined.py (Code availability). **(b**) Comparison between the median *T*_1_ estimates from Look-Locker versus Mixed within different CSF regions. The size of the markers reflects the number of voxels in the given region. Voxel region markers in the CSF mask correspond to the label of the nearest region of the FreeSurfer segmentation aseg+aparc.

### Concentrations

Studies reporting quantitative concentration estimates following intrathecal injection of gadobutrol in humans are scarce, which precludes a comparison of our estimated concentration values to a body of literature values. Table 1 and Fig. 12 shows the estimated total tracer amount in the brain and CSF in the head for each session. The tracer initially spreads through the CSF-filled spaces with 78.5% of the tracer located in the CSF after 4 h, but after 48 h and 70 h it is spread more evenly. The largest estimated amount of tracer in the head is 11.8 × 10^−2^ mmol after 24 h, corresponding to 47.2% of the injected total, and the tracer is distributed with 5.19 × 10^−2^ mmol in the brain and 6.57 × 10^−2^ mmol in the CSF.

**Figure 12.**
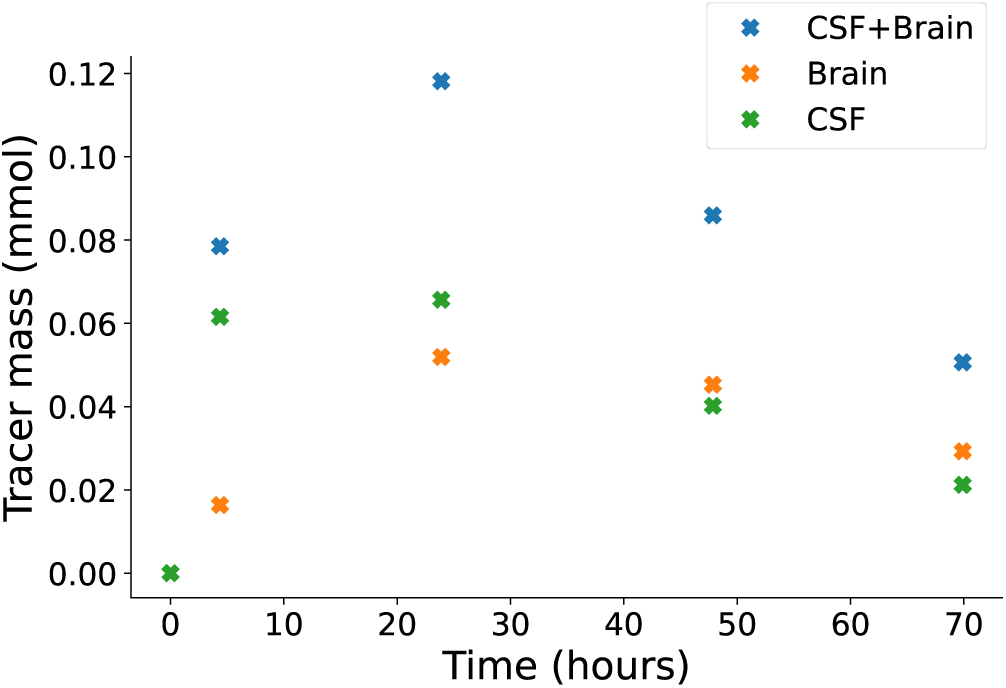
Estimated tracer amount in the brain. The brain is gray in the right figure; CSF is blue. The graph shows the total estimated tracer amount per region for each session.

**Table 1.** Total tracer amount measured within brain tissue and in the CSF-filled spaces surrounding the brain, as indicated by Fig. 12.

| Time (hours) | Tracer amount ($\times 10^{-2}$ mmol) | | |
| --- | --- | --- | --- |
|  | Brain+CSF | Brain | CSF |
| 4 | 7.85 | 1.64 | 6.16 |
| 24 | 11.8 | 5.19 | 6.57 |
| 48 | 8.59 | 4.53 | 4.02 |
| 70 | 5.06 | 2.93 | 2.12 |

There are several sources of errors with respect to quantitative concentration estimates. The uncertainty related to longitudinal relaxivity, *r*_1_, introduces a source of error in the concentration estimates. Relaxivity depends on the solvent, so we have used a value measured for water at 37 ^◦^C of 3.2 Ls^−1^ mmol^−143^, with similar values of 3.3 Ls^−1^ mmol^−1^ found in^54^. We could not find the corresponding value measured for CSF, consisting of 99% water^55^, but we note that the longitudinal relaxivity for blood plasma, with 92% water,^55^ was measured at 4.5 Ls^−1^ mmol^−1^^56^.

Moreover, we note that the concentration estimates contain negative concentrations. These can be mainly attributed to errors in the *T*_1_ maps, originating from image noise or partial volume effects, particularly at the interface between media with significantly different *T*_1_ times.

Mapping the concentration data from the MRI data voxel representation to the computational mesh introduces additional errors. MRI data may be interpreted as a piecewise-constant function on a regular Cartesian grid, which cannot be represented exactly as a piecewise-linear function on a tetrahedral mesh. As an illustration, Figure 13 shows the original concentration data at 24 h compared to the concentration after first mapping the data to the mesh and then mapping them back.

**Figure 13.**
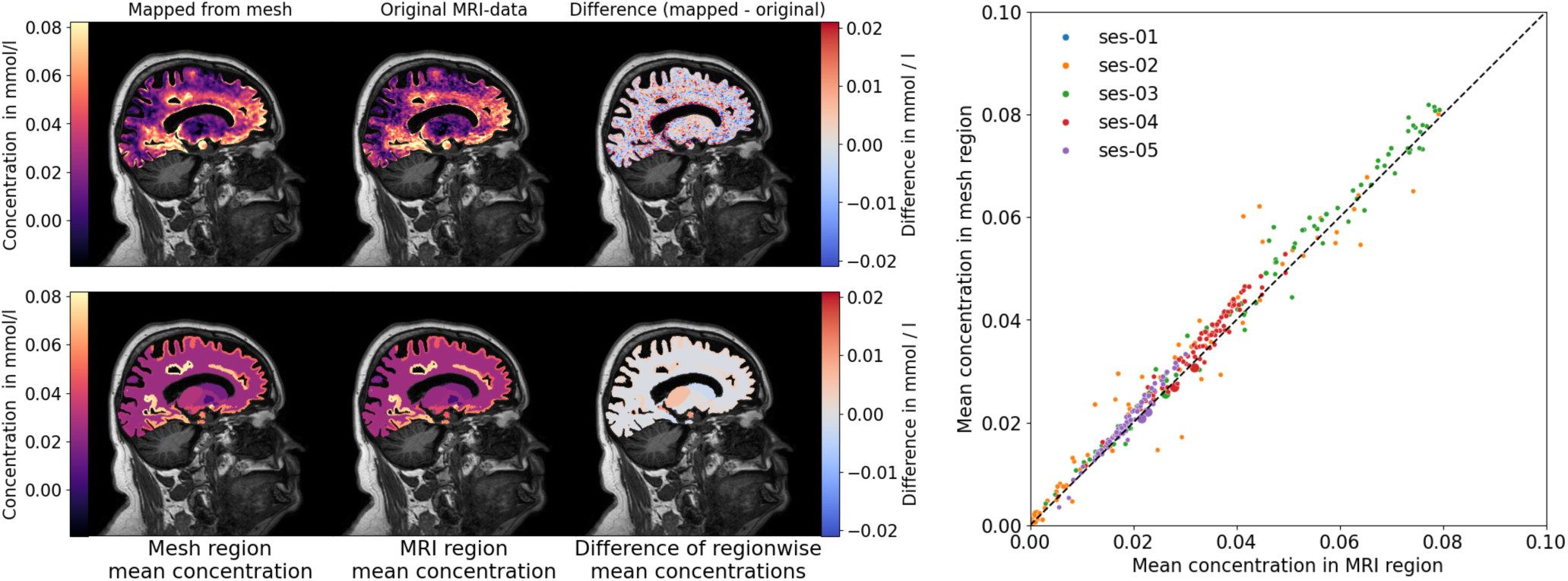
**Upper left:**Visualization of the error introduced by mapping the concentration data at 24 hours from the Cartesian MRI data grid in clinical resolution to the 3D volume mesh and back to the MR image grid. The leftmost panel shows the concentration data after mapping one roundtrip. The center panel shows the original MRI data. The rightmost panel shows the difference between the two images. The left color scale ranges from the 1^st^ to the 95^th^ percentile of the original concentration data (full range [−0.517, 0.741]). The right color scale extends to ±90^th^ percentile of the magnitude of the errors (full range of errors [−1.10, 0.80]). Negative concentrations result from noise in image data propagated through the concentration reconstruction algorithm described in Methods. **Lower left:** Alternative visualization of the error introduced by the mapping from MRI image grid to the volume mesh from the 24h session. The leftmost panel shows the MRI image grids’ mean concentration within each of the regions defined by the segmentation shown in Fig. 8d. The middle panel shown the mean concentration of each of the corresponding regions on the mesh, as shown in Fig. 8f. The third panel shows the difference between the mean concentrations. Range of color scale is shared with upper left figure. **Right:** Scatterplot comparing the regionwise mean concentrations in the MRI image with the corresponding regionwise mean concentrations in the computational mesh, with regions defined by the segmentation shown in Fig. 8d – Fig. 8f, colored by session.

### Diffusion tensor imaging (DTI) data

The mean diffusivity (MD) values for white matter and gray matter are found to be 0.77 ± 0.17mm^2^*/*s and 1.05 ± 0.17mm^2^*/*s, respectively. Due to the low resolution of the DTI, the mean diffusivity of voxels labeled as gray matter is likely to be influenced by their surroundings due to partial volume effects. Table 2 lists previously reported values of the apparent diffusion coefficients (ADC) in human brain tissue. Note that the standard deviation reported for our estimates is not comparable to those reported in the cited literature since they are reporting deviation between mean values for different subjects, whereas we report within-subject variation.

**Table 2.** *T*_1_ longitudinal relaxation times and apparent diffusion coefficient (ADC) values as reported by various sources for cerebrospinal fluid, gray matter, and white matter, compared to the values obtained in this study.

|  | cerebrospinal fluid | gray matter | white matter | reference |
| --- | --- | --- | --- | --- |
| $T_1$ in ms (mean ± std.) | 4465 ± 154<br>4163 ± 263<br>3817 ± 424<br>4391 ± 545<br>-<br>- | 1622 ± 558<br>1445 ± 119<br>1135 ± 79<br>1460 ± 33<br>1615 ± 149<br>1600 ± 110 | 955 ± 180<br>791 ± 27<br>732 ± 56<br>943 ± 57<br>911 ± 15<br>840 ± 50 | This study<br>64<br>65*<br>66<br>67<br>68 |
| ADC in $1 \times 10^{-9} \text{ ms}^{-2}$ (mean ± std.) | -<br>-<br>-<br>-<br>-<br>- | 1.05 ± 0.17<br>0.89 ± 0.04<br>0.92 ± 0.02<br>0.681 ± 0.07<br>0.67–0.83<br>- | 0.77 ± 0.17<br>0.75 ± 0.03<br>0.87 ± 0.02<br>0.613 ± 0.07<br>0.84 ± 0.11<br>0.84 ± 0.11 | This study<br>69<br>70<br>71<br>72<br>73 |

\* White matter and gray matter values are averaged over smaller reference regions used in the article.

**Table 3.** Molar concentration of gadobutrol in blood plasma, *c*_P_, in mmol L^−1^ using a direct sampling method. Reported acquisition time *t* is in minutes/hours relative to the injection time *t* = 0 min.

| $t$ | 100/1.67 | 220/3.67 | 380/4.67 | 525/8.75 | 750/12.50 | 1230/20.50 | 1515/25.25 | 2430/40.50 | 2860/47.67 |
| --- | --- | --- | --- | --- | --- | --- | --- | --- | --- |
| $c_p$ | 2.74E-04 | 4.37E-04 | 1.55E-03 | 2.07E-03 | 2.14E-03 | 2.20E-03 | 1.28E-03 | 2.31E-03 | 5.67E-04 |

**Table 4.**
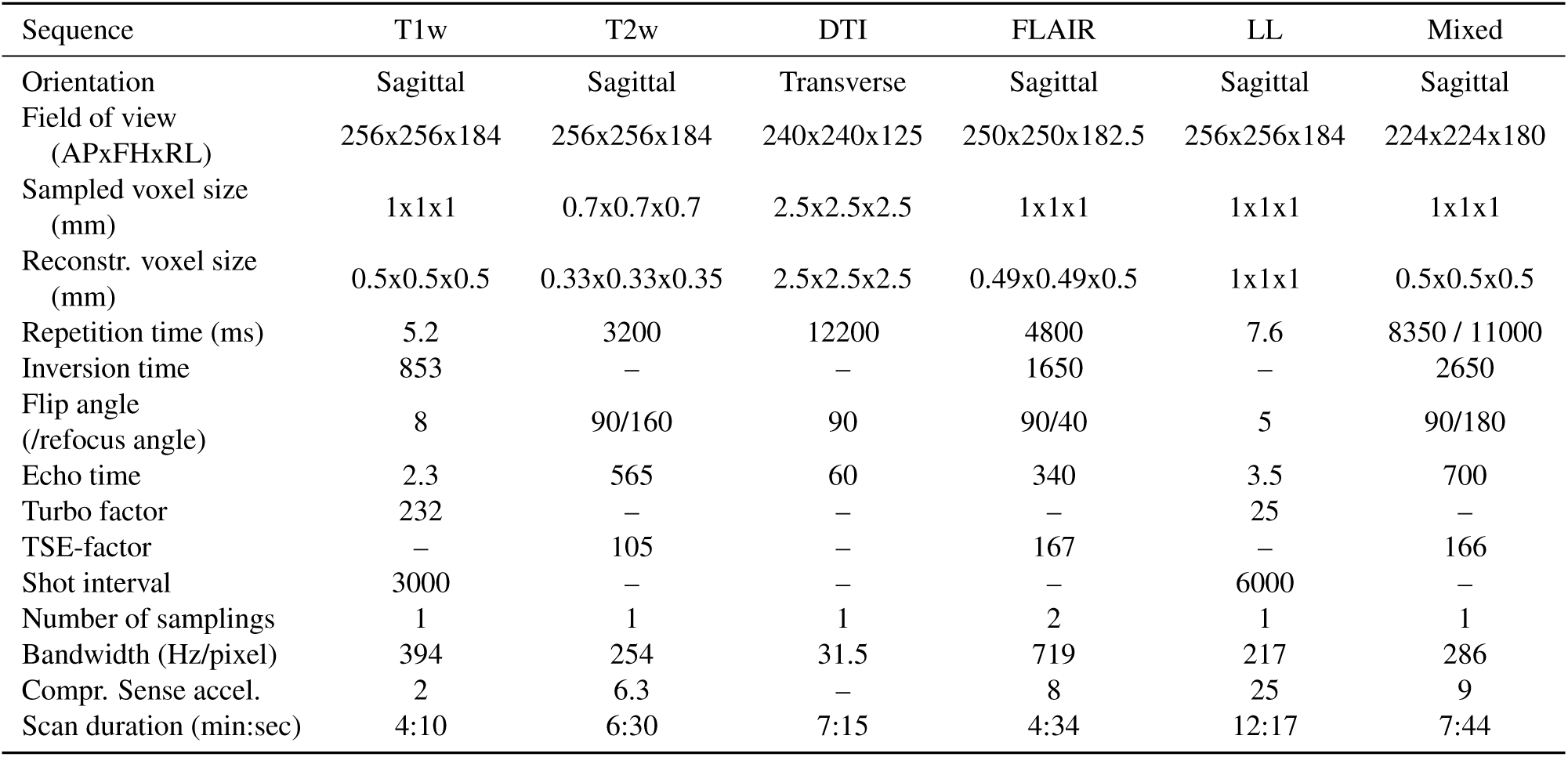
MRI sequence parameters used for the acquisition of *T*_1_-weighted images (T1w), *T*_2_-weighted images (T2w), diffusion tensor images (DTI), FLAIR, Look-locker (LL), and Mixed spin-echo inversion recovery (Mixed) images.

| Sequence | T1w | T2w | DTI | FLAIR | LL | Mixed |
| --- | --- | --- | --- | --- | --- | --- |
| Orientation | Sagittal | Sagittal | Transverse | Sagittal | Sagittal | Sagittal |
| Field of view (APxFHxRL) | 256x256x184 | 256x256x184 | 240x240x125 | 250x250x182.5 | 256x256x184 | 224x224x180 |
| Sampled voxel size (mm) | 1x1x1 | 0.7x0.7x0.7 | 2.5x2.5x2.5 | 1x1x1 | 1x1x1 | 1x1x1 |
| Reconstr. voxel size (mm) | 0.5x0.5x0.5 | 0.33x0.33x0.35 | 2.5x2.5x2.5 | 0.49x0.49x0.5 | 1x1x1 | 0.5x0.5x0.5 |
| Repetition time (ms) | 5.2 | 3200 | 12200 | 4800 | 7.6 | 8350 / 11000 |
| Inversion time | 853 | – | – | 1650 | – | 2650 |
| Flip angle (/refocus angle) | 8 | 90/160 | 90 | 90/40 | 5 | 90/180 |
| Echo time | 2.3 | 565 | 60 | 340 | 3.5 | 700 |
| Turbo factor | 232 | – | – | – | 25 | – |
| TSE-factor | – | 105 | – | 167 | – | 166 |
| Shot interval | 3000 | – | – | – | 6000 | – |
| Number of samplings | 1 | 1 | 1 | 2 | 1 | 1 |
| Bandwidth (Hz/pixel) | 394 | 254 | 31.5 | 719 | 217 | 286 |
| Compr. Sense accel. | 2 | 6.3 | – | 8 | 25 | 9 |
| Scan duration (min:sec) | 4:10 | 6:30 | 7:15 | 4:34 | 12:17 | 7:44 |

## Usage Notes

The MRI data in NIfTI format can be viewed and analyzed with a variety of open source software solutions such as FreeSurfer’s freeview, PyVista, ParaView (after unzipping). Post-processing tools used to post-process data, as described in this article, are publicly available (see Code availability). The workflow is based on Python. In addition to the description in this article, many procedures (and in parts alternative procedures) are described in^48^. In the following usage example, we show how to reuse (a) CSF concentration data as boundary data for a tracer transport simulation, (b) the brain mesh as computational mesh, and (c) DTI data as a parameter field; we then simulate tracer transport, (d) map the result onto the MRI reference image, and (e) visually compare it with concentration maps estimated from MRI. For the following usage example, we built a C++-based simulation application using the software framework DuMu^x^/DUNE framework^57–59^ with grid input and output handling via the GridFormat library^60^. As the data is provided in open standard data formats, reuse with other software should be possible analogously; we have, for instance, successfully tested a reuse scenario with the FEniCS finite element framework^48^.

### Usage example: Tracer transport simulation

Once the data has been mapped onto the mesh, it may be used in supporting computational models for solute transport in the brain, either as a target for validation or as subject-specific realistic inputs. As a usage example, we model tracer transport in the brain tissue over 72 h. The total gadolinium concentration, *c*(*x, t*) (amount per brain volume in mmol L^−1^) at position *x*, and time *t* ∈ [0, 259200] (in s), satisfies

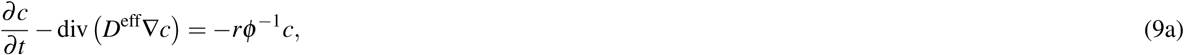

subject to the boundary and initial conditions,

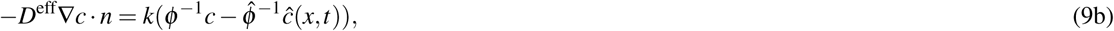

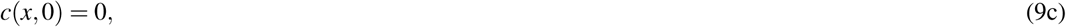

where *D*^eff^ is the effective diffusion tensor of gadolinium (in mm^2^ s^−1^), *r* is a local clearance rate (in s^−1^) due to tracer clearance to blood, *k* is the brain surface conductivity (in mms^−1^) and *ĉ*(*x, t*) is the solute concentration in the CSF just outside of the pial surface of the brain, and *φ* is the extra-cellular volume fraction of the brain tissue which is occupied by interstitial fluid. The fluid available volume fraction in the subarachnoid space (SAS) is denoted by 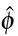. We assume that only the extra-cellular space is accessible to the tracer. Dividing the total concentration by *φ* (in tissue) or 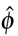 (in SAS) computes the concentration per interstitial fluid volume.

We take the values for *D*^eff^ from the provided data set data.vtu. Since the CSF concentration is only available at five discrete time points, we compute a smooth boundary data function *ĉ*(*t*) = *at* exp(−*bt*), for each degree of freedom on the boundary, fitting the parameters *a* and *b* by minimizing the discrete *l*_2_-error between *ĉ*(*t*) and the data points *Ĉ*(*t*) using a Levenberg–Marquardt algorithm. Finally, we set *φ* = 0.2, 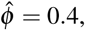 *k* = 2 × 10^−4^ mms^−1^, *r* = 1 × 10^−6^ s^−1^. A comparison of the simulation field *c*(*x, t*) and the corresponding field provided in the data set at five consecutive time points is shown in Fig. 14.

**Figure 14.**
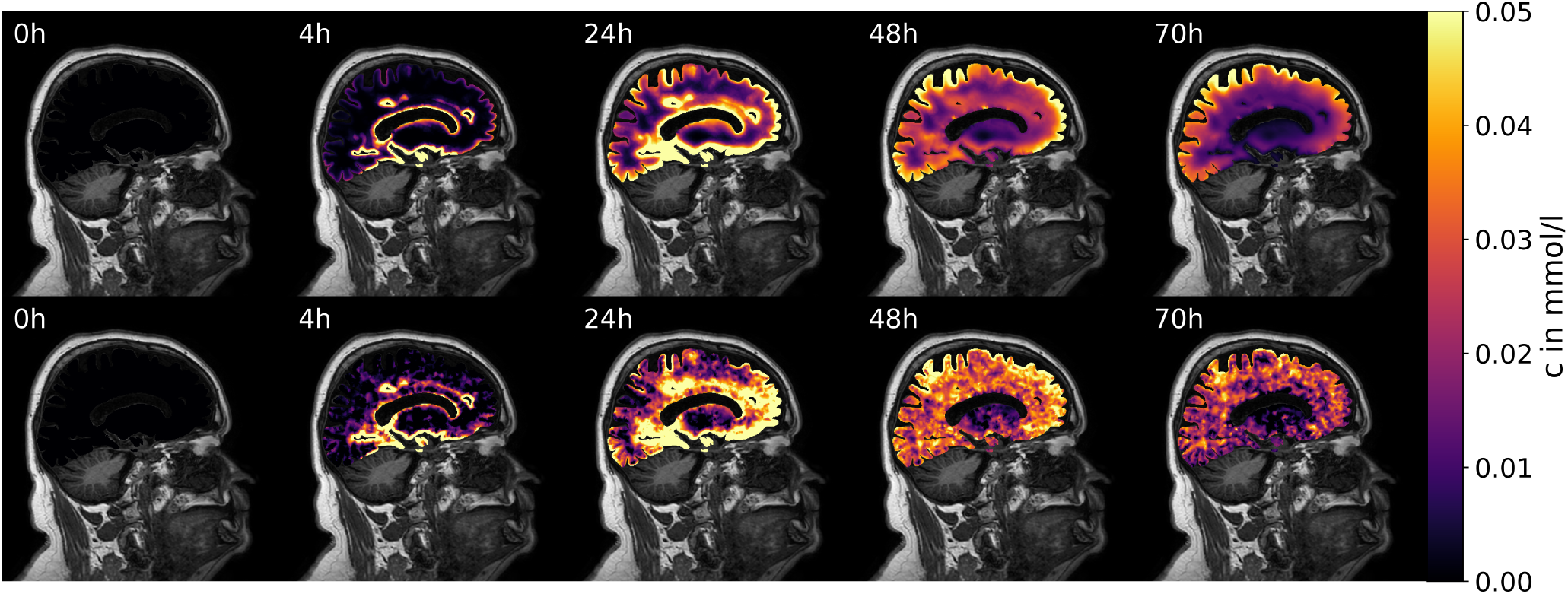
Concentration simulations. The CSF concentration data is used as the boundary condition for a finite volume-based solver of the diffusion equation. The predicted concentration within the brain tissue is compared to the concentration estimated from MRI data: (top row) simulation results; (bottom row) MRI concentration maps.

## Data Availability

All data produced are available online at https://doi.org/10.5281/zenodo.14266867

## Code availability

The source code for running each step of the described data processing pipeline is split into three repositories. The separation of the source code intends to facilitate the use of the software in future studies involving only parts of the processing pipeline of this dataset.

**gonzo:** The main repository related to this study. It includes instructions for installing necessary dependencies, running the data processing pipeline, and running scripts for creating plots in this document. The code is publicly available at https://github.com/jorgenriseth/gonzo and as archived dataset^61^.

**gMRI2FEM:** A Python library used for post-processing MRI data. The code is publicly available at https://github. com/jorgenriseth/gMRI2FEM and as archived dataset^62^.

**dumux-braindiffusion-miniapp:** A reuse example code that provides a simulator used in the Usage example and for Fig. 14 (C++ code, based on the DuMu^x^/DUNE framework^57–59^ and GridFormat^60^). The code is publicly available at https://github.com/timokoch/dumux-braindiffusion-miniapp and as archived dataset^63^.

## Acknowledgements

The authors thank Geir Ringstadt and Per-Kristian Eide for spearheading the development of glymphatic MRI and for fruitful discussions. The authors are grateful to the anonymous volunteer for participating in the study and giving informed consent to open publication, allowing us to disseminate this unique data set.

S.L. and K.N. acknowledge funding from the South Eastern Norway Health Authority (Helse Sør-Øst) within project 2022022 (Clearance pathways in Parkinson’s disease) and the Norwegian Health Association (Nasjonalforeningen for folkehelsen) within projects 25598 and 28398. T.K. acknowledges funding from the European Union’s Horizon 2020 Research and Innovation programme under the Marie Skłodowska-Curie Actions Grant agreement No 801133 (Scientia fellows II). T.K. and K.A.M. acknowledge funding by the Research Council of Norway, project 301013 (Alzheimer’s physics). T.K., J.N.R and K.A.M. acknowledge funding by the European Research Council under grant 101141807 (aCleanBrain). K.A.M. and L.M.W acknowledges the funding from the “Computational Hydrology project” a strategic Sustainability initiative at the Faculty of Natural Sciences, UiO. K.A.M. acknowledges funding from the Stiftelsen Kristian Gerhard Jebsen via the K. G. Jebsen Centre for Brain Fluid Research and the national infrastructure for computational science in Norway, Sigma2, via grant NN9279K. The work of L.T.Z. was supported in part by U.S. NSF DMS-220829 and the U.S.-Norway Fulbright Foundation.

## Author contributions statement

J.R.: Data curation, Formal analysis, Investigation, Methodology, Software, Validation, Visualization, Writing – original draft; T.K.: Formal analysis, Investigation, Software, Methodology, Supervision, Validation, Visualization, Writing – original draft; S.L.: Data curation, Methodology, Investigation, Writing – review & editing; T.H.S.: Data curation, Formal analysis, Methodology, Writing – review & editing; L.M.V.: Software, Methodology, Writing – review & editing; L.Z.: Conceptualization, Investigation, Writing – review & editing; K.N.: Conceptualization, Funding acquisition, Methodology, Project Administration, Supervision, Writing – review & editing; K.A.M.: Conceptualization, Methodology, Funding acquisition, Project Administration, Supervision, Writing – original draft;

## Footnotes

1 The presence of contrast agent increases both the longitudinal relaxation rate *R*_1_= 1*/T*_1_ and the transversal relaxation rate *R*_2_ = 1*/T*_2_ (that is, it shortens the relaxation times *T*_1_ and *T*_2_) of the sample region by an amount proportional to its concentration; also see Section Methods, Eq. (7).

2 The data is sparse in time, sparse in space in the sense that only imaging data of the head is provided while the injection site is in the lumbar region, and sparse on a group-level since it contains data from a single subject.

3 https://github.com/rordenlab/dcm2niix^35^

4 https://github.com/SVMTK/SVMTK

5 https://crai.no/product/nordicice

